# Multimorbidity and Frailty in Older Adults: A Comparative Study of Care Models and Policy Strategies in Spain, Italy, Chile, and Colombia

**DOI:** 10.64898/2026.07.25.26358906

**Authors:** Omaira Valencia, Sebastián Leon-Giraldo, Clara Donnoli, Manuel Miron-Rubio, Paula Zamorano, Nicola Pinelli, Giuseppe Liotta, Luis Fernando Gutierrez-Fernandez, Oscar Bernal

## Abstract

**Background:** Population aging is rapidly increasing the burden of multimorbidity and frailty, yet there is limited evidence on how healthy aging policies are implemented across different demographic and health system contexts.

**Methods:** We conducted a mixed-methods comparative case study in Spain, Italy, Chile, and Colombia, integrating a Targeted review of policy documents and relevant literature, quantitative indicators, and expert interviews.

**Results:** Chile and Colombia showed faster aging dynamics (compound annual growth rates of 4.18% and 4.21%) than Italy (1.96%) and Spain (1.39%), with estimated doubling times of 17 versus 36–50 years. All four countries have established policy frameworks; implementation remains uneven. Key barriers included fragmented services, limited community care, workforce constraints, and misaligned financing.

**Conclusions:** The central challenge lies not in the absence of policy frameworks but in their effective implementation. Strengthening primary care, embedding frailty assessment, and improving health–social care integration are key priorities for systems facing rapid demographic transition.

## Introduction

Population aging represents one of the defining demographic transformations of the twenty-first century, with profound consequences for population health and the organization of health and social care systems. Rising life expectancy has been accompanied by a growing burden of age-related conditions—most notably multimorbidity and frailty—which frequently coexist and interact, compounding vulnerability and care complexity among older adults (Barnett et al., 2012; Marengoni et al., 2011). Frailty is understood as a clinical state of heightened vulnerability resulting from cumulative decline across multiple physiological systems, associated with increased risks of disability, hospitalisation, institutionalization, and mortality (Clegg et al., 2013; Hoogendijk et al., 2019). The 2025 Lancet Commission on Frailty has further underscored frailty as the foremost geriatric syndrome of our time, noting that without adequate understanding of its causal pathways, policy-makers cannot develop effective preventive strategies (Clegg et al., 2025). When combined with multimorbidity, frailty further complicates clinical management and amplifies the demand for coordinated, long-term care.

These demographic and epidemiological trends challenge traditional disease-oriented models of health care delivery. There is growing recognition that care for older adults must move beyond single-disease approaches towards integrated, person-centerd models that address functional capacity and complex care needs (World Health Organization [WHO], 2015; Beard et al., 2016; Cesari et al., 2018). International frameworks on healthy aging emphasize the maintenance of functional ability, the strengthening of primary and community care, and the integration of health and social services as core responses to the needs of aging populations (Beard et al., 2016; Dent et al., 2019).

Although multimorbidity and frailty are global phenomena, their prevalence and implications vary according to demographic trajectories, socioeconomic conditions, and health system organization (Salisbury et al., 2018). Countries at advanced stages of demographic aging, particularly in Southern Europe, already confront high levels of multimorbidity and long-term care demand. By contrast, many middle-income countries are undergoing accelerated aging within health systems that remain fragmented and unevenly resourced (Briggs et al., 2018; Lloyd-Sherlock et al., 2016). In Latin America, this combination of rapid demographic change and institutional constraints creates particular challenges for the development of integrated responses to complex needs in later life (Gutiérrez Robledo et al., 2022; Dlima et al., 2024). Recent evidence from Nature Medicine further indicates that social disparities, cardiometabolic disease, and mental health are the principal predictors of healthy aging in Latin American populations—factors more pronounced in lower-income countries and qualitatively distinct from patterns documented in Europe (Ibanez et al., 2023).

Comparative analyzes across countries at different stages of demographic transition offer valuable insights into how health systems adapt to multimorbidity and frailty. Such comparisons illuminate shared barriers, emerging care models, and policy strategies that may inform the development of more coordinated and sustainable responses (Orueta et al., 2013). This study compares four countries as national units of analysis to examine how policy frameworks on healthy aging are translated into implementation, with particular attention to screening approaches and health-social care integration, in order to assess system preparedness for responding to population aging, multimorbidity, and frailty.

## Materials and Methods

### Study Design

We used a convergent mixed-methods comparative case study design to assess country-level preparedness to respond to multimorbidity and frailty among older adults in Italy, Spain, Chile, and Colombia. Countries were treated as the primary unit of analysis and compared across three analytical dimensions: policy implementation, screening and identification capacity, and health-social care integration. A mixed-methods approach was appropriate because these dimensions involve both measurable system characteristics and implementation processes that cannot be adequately understood through a single method alone. Quantitative indicators were used to characterize demographic aging and health system context, policy and document review to identify formal commitments and institutional arrangements, and qualitative interviews and field-based observations to examine how these policies and models were operationalized in practice, including barriers, gaps, and contextual adaptations.

### Comparative framework

The comparative framework was organized around three dimensions: policy implementation, screening and identification capacity, and health-social care integration because together they capture whether national systems are operationally prepared to respond to population aging beyond the formal existence of policy documents. Policy implementation reflects the extent to which formal commitments have been translated into programs and service delivery arrangements; screening and identification capacity reflects the ability of systems to detect frailty, multimorbidity, and complex care needs; and health-social care integration reflects the extent to which responses to older adult needs are coordinated across sectors.

### Case selection

The four countries were purposively selected as analytically contrasting cases rather than as representative examples of all OECD members. Selection aimed to capture variation in the pace of demographic aging and in the development of policy and service responses to older adult care needs, while maintaining sufficient comparability in terms of data availability and institutional frameworks. Italy and Spain represent more advanced aging contexts with longer-standing policy experience, whereas Chile and Colombia provide insight into Latin American settings facing accelerated aging and evolving system responses. This configuration allowed us to examine both shared implementation challenges and the potential transferability of lessons across countries at different stages of demographic transition.

### Data Collection

We collected data from four complementary sources, each capturing distinct dimensions of the phenomenon under study: demographic trends, epidemiological patterns, health system capacity, and implementation dynamics.

#### Quantitative secondary data

We compiled country-level quantitative data to characterize epidemiological profiles, demographic aging trajectories, and health system capacity in each country. Sources included national statistical offices, ministries of health, and international databases (OECD, World Bank, WHO). Indicators covered population age structure, prevalence of selected chronic conditions, hospital bed density, utilisation of hospital and primary care services, and long-term care (LTC) resources. These data provided a standardized quantitative context for cross-country comparison of system readiness and service pressure associated with aging, multimorbidity, and frailty.

#### Targeted review of policy documents and relevant literature

A targeted review of policy documents and relevant literature was conducted to identify national policy frameworks, institutional arrangements, and programmatic responses related to multimorbidity, frailty, and healthy aging in the four study countries. The review also served to contextualize country differences and to inform the comparative analytical framework used in the study. Searches were undertaken in PubMed, Scopus, and Google Scholar, complemented by targeted searches of official government, ministry of health, and international organization websites. Search terms combined concepts related to older adults, aging, multimorbidity, frailty, integrated care, healthy aging, long-term care, and country-specific policy responses.

Documents were eligible for inclusion if they addressed national or subnational policies, programs, care models, or system responses relevant to older adults aged 60 years and over, particularly in relation to frailty, multimorbidity, prevention, long-term care, or health and social care coordination. Peer-reviewed articles, policy reports, strategic plans, and official institutional documents published in English, Spanish or Italian between 2010 and January 2026 were considered. Documents focused exclusively on younger populations, single-disease management without relevance to broader aging-related complexity, or highly local initiatives with no policy or system relevance were excluded.

Rather than aiming for exhaustive systematic coverage, this targeted review was designed to identify the most relevant policy and literature sources needed to characterize formal policy commitments, service organization, and implementation approaches across countries. Findings from the review were used to support cross-country comparison and to contextualize the interpretation of quantitative and qualitative evidence.

#### Expert interviews

The qualitative component consisted of in-depth expert interviews with a purposive sample of senior informants selected for their direct knowledge of aging policy, service delivery, and implementation processes in the four countries. The purpose of this component was not to achieve broad stakeholder representation, but to generate focused comparative insight into how national responses to multimorbidity and frailty among older adults were being translated into practice, and to identify key implementation barriers, system gaps, and cross-country differences in preparedness.

Six experts were recruited through purposive sampling, prioritizing individuals with recognized experience in older adult health, geriatrics, health system organization, integrated care, or policy implementation. Interviews were conducted using a semi-structured interview guide organized around five domains: (1) multimorbidity and care models; (2) frailty measurement tools and their clinical and policy use; (3) evaluation, monitoring, and perceived impact; (4) governance and sustainability; and (5) lessons learned and recommendations (Supplementary Material 1). These domains were designed to elicit expert perspectives on how formal policy commitments had been operationalized in practice and to identify transferable lessons across countries.

Interviews were conducted virtually between April and May 2025, lasted approximately 45–60 minutes, all participants provided verbal informed consent before participation. Consent to participate and to audio recording was obtained at the beginning of each Microsoft Teams interview and documented through the meeting recording. Audio files were transcribed and de-identified prior to analysis. Transcripts were analyzed thematically using NVivo, with coding guided by the study’s comparative framework and focused on recurrent themes related to policy implementation, institutional capacity, coordination challenges, and policy-to-practice gaps. The interview findings were used to complement and interpret the quantitative and documentary evidence, rather than to support broad representational claims.

### Data Analysis

#### Quantitative analysis

Country-level quantitative descriptors of the sociodemographic and health system context for Italy, Spain, Chile, and Colombia were compiled from policy documents and authoritative statistical repositories, including the World Bank Health, Nutrition and Population Statistics and OECD databases (latest available editions). For each indicator, the most recent available year and the longest historical series (typically spanning 2000–2024) were extracted. Indicators were reported using standardized units as provided by the original sources. No hypothesis testing or model-based analyses were performed; missing or non-reported values were not imputed and are explicitly indicated as “not available.”

#### Demographic transition assessment

Demographic transition velocity was assessed using aging index calculations derived from OECD Historical Population Data (DSD_POPULATION@DF_POP_HIST) spanning 2000–2024 (OECD, 2024). The aging index was calculated as the ratio of the population aged ≥65 years to the population aged 0–14 years, multiplied by 100, following the methodology of the United Nations Population Division (United Nations, 2022). Compound Annual Growth Rate (CAGR) was computed for three distinct periods (2000–2010, 2010–2020, 2020–2024) using the formula: CAGR = [(Ending Value/Beginning Value)^(1/n) − 1] × 100. Population doubling time was estimated using the standard Rule of 70 approximation: Doubling Time = 70 / Overall CAGR.

#### Qualitative analysis

Expert interview data were transcribed, de-identified, and analyzed thematically in NVivo using a hybrid deductive–inductive approach. An initial coding framework was developed from the interview guide and the study’s comparative framework, including domains related to multimorbidity and care models, frailty measurement and use, evaluation and perceived impact, governance and sustainability, and lessons learned for implementation. This framework was refined iteratively as additional themes emerged from the data. Two researchers reviewed the transcripts and coding structure, with discrepancies discussed and resolved through consensus to strengthen analytic consistency. Analytic decisions were documented through memos, coding notes, and versioned codebooks to enhance transparency and rigor. The qualitative analysis was used to identify recurrent themes related to policy implementation, institutional capacity, coordination challenges, and policy-to-practice gaps, and to support cross-country interpretation of differences in system preparedness.

#### Cross-country synthesis

The cross-country synthesis was conducted through a structured and iterative comparison of findings across countries and data sources. Evidence from the targeted policy and literature review, quantitative indicators, and expert interviews was organized within a thematic matrix aligned with the study’s predefined analytic domains of policy implementation, screening and identification capacity, and health-social care integration. This process enabled the systematic identification of convergent and divergent patterns across countries and supported interpretation of how demographic context, institutional arrangements, and implementation practices shape national responses to multimorbidity and frailty. The synthesis did not aim for statistical generalization; rather, it was intended to identify shared challenges, context-specific approaches, and lessons of potential relevance for countries at different stages of demographic transition.

### Ethical Considerations

Ethical approval was granted by the Research Ethics Committee of the Fundación Santa Fe de Bogotá (approval communication: CCEI-16596-2024, issued 27 June 2024). All expert interview participants provided verbal informed consent prior to participation. Interviews were conducted virtually, and all participants agreed to participate and to be audio-recorded.

## Results

### Demographic and Epidemiological Context of Aging Across Countries

Table 1 and Figure 1 present aging index trajectories for the four countries between 2000 and 2024. Italy and Spain entered the study period with aging indices of 127.9 and 113.2 per 100 persons, respectively, whereas Chile (28.0) and Colombia (17.5) had substantially lower baseline values. By 2024, Italy reached 203.7, Spain 157.8, Chile 74.9, and Colombia 47.1, corresponding to percentage increases of 59.3%, 39.4%, 167.5%, and 169.1%, respectively (Table 1).

**Figure 1.**
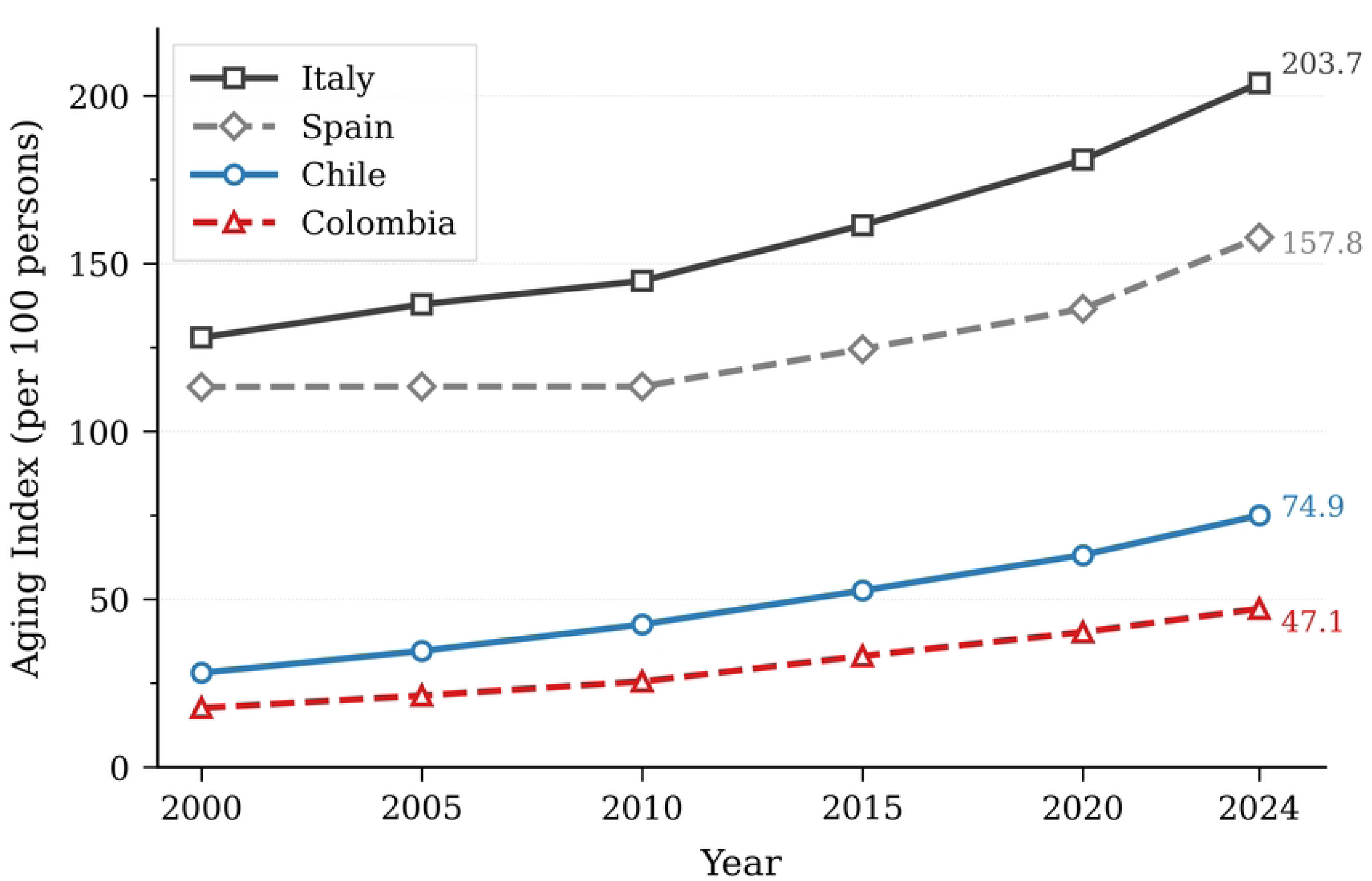
Temporal Trends in Population Aging Index, Chile, Colombia, Italy, and Spain (2000–2024)

**Table 1.** Comparative Analysis of Aging Index Dynamics in Chile, Colombia, Italy, and Spain (2000-2024)

| Country | Aging Index Values |  | Compound Annual Growth Rate (%) |  | Summary Metrics |  |  |  |  |  |  |
| --- | --- | --- | --- | --- | --- | --- | --- | --- | --- | --- | --- |
|  | 2000 | 2010 | 2020 | 2024 | 2000-10 | 2010-20 | 2020-24 | Overall | DT (years) | Δ | % Inc |
| Chile | 28.0 | 42.4 | 63.1 | 74.9 | 4.24 | 4.06 | 4.38 | 4.18 | 16.7 | 46.9 | 167.5 |
| Colombia | 17.5 | 25.4 | 40.1 | 47.1 | 3.80 | 4.67 | 4.10 | 4.21 | 16.6 | 29.6 | 169.1 |
| **Italy** | 127.9 | 144.8 | 180.9 | 203.7 | 1.25 | 2.25 | 3.01 | 1.96 | 35.7 | 75.8 | 59.3 |
| **Spain** | 113.2 | 113.3 | 136.5 | 157.8 | 0.01 | 1.88 | 3.69 | 1.39 | 50.2 | 44.6 | 39.4 |

*Note: The aging index represents the number of persons aged ≥65 years per 100 persons aged 0-14 years. Values shown for 2000, 2010, 2020, and 2024. CAGR (Compound Annual Growth Rate) calculated as $[(\text{Ending Value}/\text{Beginning Value})^{(1/n)} - 1] \times 100$ , where $n$ = number of years in the period. DT (Doubling Time) estimated using the Rule of 70 ( $70/\text{Overall CAGR}$ ), indicating years required to double the aging index at current growth rates. $\Delta$ = Absolute change in aging index from 2000 to 2024. % Inc = Percent increase from baseline (2000) to 2024.*
*Tan-shaded rows indicate Latin American countries; blue-shaded rows indicate Southern European countries. Bold values in the 'Overall' column highlight the average annual growth rate across the entire study period (2000-2024), the primary metric for cross-country comparison.*
*Data source: Authors' calculations based on Organisation for Economic Co-operation and Development (OECD). Historical Population Data (Dataset: DSD\_POPULATION@DF\_POP\_HIST). Values for 2000-2023 represent observed data; 2024 values are demographic projections. Paris: OECD Publishing; 2024. Available from: https://data-explorer.oecd.org*

Compound Annual Growth Rates differed markedly between the two country groups. Chile and Colombia recorded overall CAGRs exceeding 4% (4.18% and 4.21%, respectively), with estimated doubling times of approximately 17 years. Italy and Spain showed overall CAGRs of 1.96% and 1.39%, with doubling times of 35.7 and 50.2 years, respectively. Period-specific growth rates revealed an accelerating trend in three of the four countries during 2020–2024: Spain increased from 1.88% to 3.69%, Italy from 2.25% to 3.01%, and Chile from 4.06% to 4.38%. Colombia, by contrast, showed a slight deceleration, from 4.67% in 2010–2020 to 4.10% in 2020– 2024, although its growth rate remained the highest among the four countries in the earlier period (Table 1).

Regarding health system capacity, avoidable mortality rates were lower in Italy (146 per 100,000) and Spain (163) than in Chile (247) and Colombia (328), compared with an OECD average of 237. Life expectancy at age 65 was 21.9 years in Spain, 21.2 in Italy, 20.7 in Chile, and 18.7 in Colombia, against an OECD average of 20.0 years. Diabetes prevalence ranged from 6.4% in Italy to 10.8% in Chile, with Spain at 10.3% and Colombia at 8.3% (OECD average: 7.0%). Spain reported 43.9 long-term care beds per 1,000 population aged 65 years and over, exceeding the OECD average of 41.2, whereas Italy reported 21.8. Unmet long-term care needs among persons with at least one ADL or IADL limitation were 34.8% in Italy and 36.2% in Spain, both below the OECD average of 47.1%. No comparable long-term care data were available for Chile or Colombia (Table 2).

**Table 2.** Health System Capacity and Preparedness Indicators, 2023 or nearest year.

| Country | Avoidable Mortality<br>(per 100,000) | Diabetes<br>Prevalence (%) | Unmet LTC Needs<br>≥1 ADL/IADL (%) | LTC Beds per<br>1,000 (≥65) | Life Expectancy<br>at 65 (years) |
| --- | --- | --- | --- | --- | --- |
| OECD Average | 237 | 7,0 | 47,1 | 41,2 | 20,0 |
| Chile | 247 | 10,8 | ND | ND | 20,7 |
| Colombia* | 328 | 8,3 | ND | ND | 18,7 |
| Italy | 146 | 6,4 | 34,8 | 21,8 | 21,2 |
| Spain | 163 | 10,3 | 36,2 | 43,9 | 21,9 |

Notes: LTC = Long-term care; ADL = Activities of Daily Living; IADL = Instrumental Activities of Daily Living; ND = No data available
\* Latest data from 2020-2022. Accession/partner country.
Data sources: OECD (2025), Health at a Glance 2025: OECD Indicators, OECD Publishing, Paris, https://doi.org/10.1787/8f9e3f9

Estimated multimorbidity prevalence among older adults varied across countries: 50–60% in Spain, 45–55% in Italy, 35–45% in Chile, and 73.1% in Colombia, although differences in age groups, data sources, and methodological approaches limit direct comparability (Supplement 2).

### National Policy and Regulatory Frameworks

A review of national policy documents and technical sources revealed that all four countries have established formal policy frameworks addressing population aging, although with differences in scope and operational detail (Table 3).

**Table 3.** Comparative Policy Frameworks and Implementation Patterns on Aging, Frailty, and Multimorbidity.

| Country | National Aging Policy Framework | Frailty and Multimorbidity in Health Policy | Care Model Orientation and Entry Points | Implementation Status | Main Sources |
| --- | --- | --- | --- | --- | --- |
| Spain | Consolidated national frameworks for aging and chronicity; dual strategy combining chronic care and healthy longevity. | Multimorbidity addressed within chronicity strategies; frailty explicitly addressed through national consensus and dedicated roadmap. Risk stratification plus Fried phenotype screening in primary care; nursing plays a central role. | Proactive, primary care–centered model with integrated care pilots. PHC-led pathways coordinating primary, hospital, and social services; early discharge and continuity of care prioritized. | Intermediate–advanced; heterogeneous across autonomous communities. Trained teams and shared screening criteria in place; constraints include high workload and underfunding. | Ministerio de Sanidad (2012, 2022); ADVANTAGE Joint Action; national consensus on frailty; expert interviews. |
| Italy | Comprehensive reform (Law 33/2023) recognizing biopsychosocial frailty as a gatekeeping criterion for care access. | Multimorbidity addressed indirectly within chronic and non-self-sufficiency policies. Frailty embedded within multidimensional assessment frameworks (biopsychosocial) linked to Integrated Individual Care Plans. | Integrated socio-health continuum, community-oriented. Community Houses integrating health and social care; PHC entry with case management and multiprofessional teams. | Early–intermediate; reform phase. Policy commitment to community-based care established; implementation largely dependent on regional planning. Limited community services and workforce constraints. | Presidenza del Consiglio dei Ministri (2023); FIASO (2024); expert interviews. |
| Chile | Long-standing national policy on older persons; nationwide Family Health Model with uneven regional implementation. | Multimorbidity explicitly included in chronic care strategies. Functional assessment used as proxy for vulnerability; frailty not operationalized as a standardized clinical construct. Gradual shift from diagnosis-led to biopsychosocial/person-centered approach. | Primary care–based, person-centered model. PHC hub with growing links to the social sector; historically diagnosis-led approach under transition. | Intermediate, pilot-based. Strong stewardship at Ministry level; pilot programs and targeted resource allocation enable progress. Ongoing cultural transition from diagnosis-centered to person-centered care. | MINSAL (2021a, 2021b); Zamorano et al. (2022); expert interviews. |
| Colombia | National Policy on Aging and Old Age (2022–2031); broad intersectoral framework focused on autonomy, functional ability, and social determinants. | Multimorbidity supported by strong diagnostic evidence (World Bank analytical studies). Frailty measurement reported in some settings using the FRAIL scale; clinical focus centered on functional capacity rather than morbidity counts. | Risk-based integrated model (conceptual). Long-term care framed as a country-level challenge; no articulated national care coordination model. | Early, pilot-based, fragmented. Progress mainly in governance and intersectoral coordination; health-sector interventions limited to pilots and subnational initiatives. System fragmentation and uneven territorial implementation. | Decreto 681/2022; World Bank (2023, 2024); Bolívar Vargas et al. (2024); Ministerio de Salud y Protección Social (2023); DNP (2024); expert interviews. |

*Note: This table integrates evidence from national policy documents, technical reports, peer-reviewed publications, and expert interviews (n = 6). "Implementation status" reflects the degree of documented operationalization rather than policy intent or measured outcomes. No causal or performance comparisons are implied. Tan-shaded rows = Latin American countries; blue-shaded rows = Southern European countries. All citations use author–year format consistent with the Journal of Aging and Health (SAGE) style.*

In Spain, national strategies on chronicity and frailty are complemented by technical consensus documents and a dedicated roadmap for the care of older adults in primary and community settings (Ministerio de Sanidad, 2012, 2022). In Italy, recent legislative reform on aging and non-self-sufficiency establishes an integrated socio-health framework for older adult care, situating the response within a continuum of health and social services and emphasizing multidimensional assessment and territorial organization of care; implementation is at an early stage and largely dependent on regional planning instruments (Presidenza del Consiglio dei Ministri, 2023; FIASO, 2024).

In Chile, multimorbidity has been incorporated into national health strategies, with emphasis on person-centered care, functional capacity, and prevention of dependency within primary care, although frailty is not defined as a standardized clinical construct within these policy documents (MINSAL, 2021a, 2021b; Zamorano et al., 2022). In Colombia, the National Policy on Aging and Old Age (2022–2031) establishes a broad intersectoral framework focused on autonomy, functional ability, and social determinants; diagnostic analyses document a high burden of multimorbidity among older adults (World Bank, 2023; Bolívar Vargas et al., 2024; Ministerio de Salud y Protección Social, 2023). Monitoring reports for the 2023 implementation period describe progress in governance and intersectoral coordination, while health-sector interventions addressing complex clinical needs remain limited (Departamento Nacional de Planeación, 2024). Across the four countries, reviewed documents described varying degrees of translation from policy frameworks into operational practice, ranging from early-stage national reforms to regionally heterogeneous pilot-based initiatives (Table 3).

### Care Models and Implementation: Expert Interview Findings

Six key informants were interviewed, representing senior clinicians, program managers, and policy analysts from Italy, Spain (Community of Madrid), Colombia, and Chile. All had a minimum of ten years of experience in designing, implementing, or evaluating services for older adults with multimorbidity or frailty.

Qualitative coding yielded 99 references distributed across four thematic categories: frailty assessment (n = 37), barriers (n = 11), enablers and success conditions (n = 11), and lessons learned (n = 27). Table 4 presents country-level findings on frailty assessment approaches, reported barriers, and enabling factors.

**Table 4.**
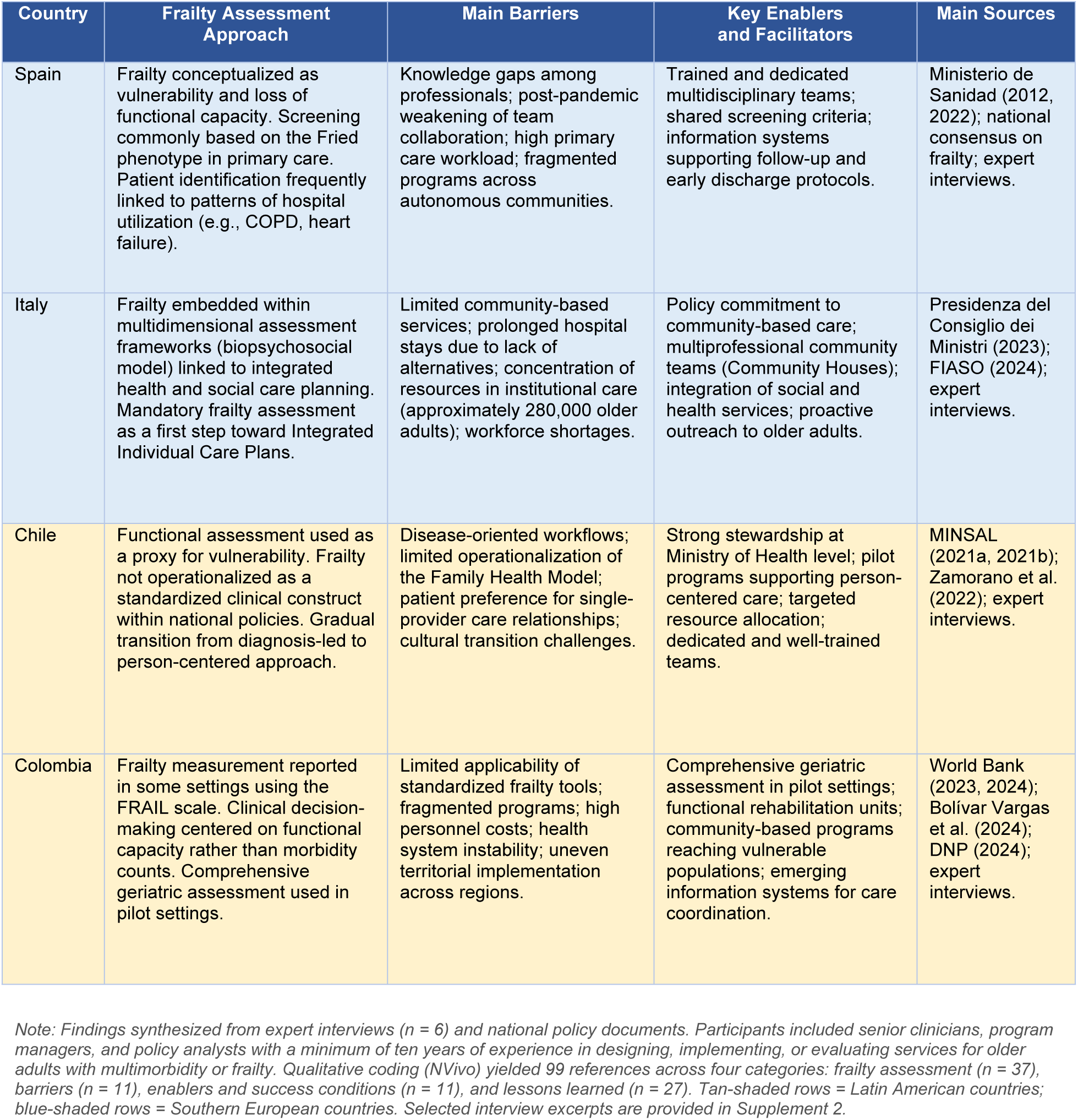
Frailty Assessment Approaches, Barriers, and Enabling Factors by Country.

| Country | Frailty Assessment Approach | Main Barriers | Key Enablers and Facilitators | Main Sources |
| --- | --- | --- | --- | --- |
| Spain | Frailty conceptualized as vulnerability and loss of functional capacity. Screening commonly based on the Fried phenotype in primary care. Patient identification frequently linked to patterns of hospital utilization (e.g., COPD, heart failure). | Knowledge gaps among professionals; post-pandemic weakening of team collaboration; high primary care workload; fragmented programs across autonomous communities. | Trained and dedicated multidisciplinary teams; shared screening criteria; information systems supporting follow-up and early discharge protocols. | Ministerio de Sanidad (2012, 2022); national consensus on frailty; expert interviews. |
| Italy | Frailty embedded within multidimensional assessment frameworks (biopsychosocial model) linked to integrated health and social care planning. Mandatory frailty assessment as a first step toward Integrated Individual Care Plans. | Limited community-based services; prolonged hospital stays due to lack of alternatives; concentration of resources in institutional care (approximately 280,000 older adults); workforce shortages. | Policy commitment to community-based care; multiprofessional community teams (Community Houses); integration of social and health services; proactive outreach to older adults. | Presidenza del Consiglio dei Ministri (2023); FIASO (2024); expert interviews. |
| Chile | Functional assessment used as a proxy for vulnerability. Frailty not operationalized as a standardized clinical construct within national policies. Gradual transition from diagnosis-led to person-centered approach. | Disease-oriented workflows; limited operationalization of the Family Health Model; patient preference for single-provider care relationships; cultural transition challenges. | Strong stewardship at Ministry of Health level; pilot programs supporting person-centered care; targeted resource allocation; dedicated and well-trained teams. | MINSAL (2021a, 2021b); Zamorano et al. (2022); expert interviews. |
| Colombia | Frailty measurement reported in some settings using the FRAIL scale. Clinical decision-making centered on functional capacity rather than morbidity counts. Comprehensive geriatric assessment used in pilot settings. | Limited applicability of standardized frailty tools; fragmented programs; high personnel costs; health system instability; uneven territorial implementation across regions. | Comprehensive geriatric assessment in pilot settings; functional rehabilitation units; community-based programs reaching vulnerable populations; emerging information systems for care coordination. | World Bank (2023, 2024); Bolívar Vargas et al. (2024); DNP (2024); expert interviews. |

*Note: Findings synthesized from expert interviews (n = 6) and national policy documents. Participants included senior clinicians, program managers, and policy analysts with a minimum of ten years of experience in designing, implementing, or evaluating services for older adults with multimorbidity or frailty. Qualitative coding (NVivo) yielded 99 references across four categories: frailty assessment (n = 37), barriers (n = 11), enablers and success conditions (n = 11), and lessons learned (n = 27). Tan-shaded rows = Latin American countries; blue-shaded rows = Southern European countries. Selected interview excerpts are provided in Supplement 2.*

In Spain, frailty was conceptualized as vulnerability and loss of functional capacity, with screening commonly based on the Fried phenotype in primary care settings; patient identification was frequently linked to patterns of hospital utilization. In Italy, frailty assessment was embedded within multidimensional evaluation frameworks connected to integrated health and social care planning. In Chile, functional assessment served as a proxy for vulnerability, without formal operationalization of frailty as a standardized clinical construct. In Colombia, frailty measurement was reported in some settings using the FRAIL scale, with clinical decision-making centered on functional capacity rather than morbidity counts.

Barriers reported across countries included knowledge gaps among professionals, weakened team collaboration, high primary care workload, limited community-based services, concentration of resources in institutional care, and uneven territorial implementation. Enabling factors highlighted by participants included trained and dedicated multidisciplinary teams, shared assessment criteria, integration of health and social services, and information systems supporting follow-up and care coordination (Table 4; Supplement 3).

## Discussion

This comparative analysis examined how four countries at different stages of demographic transition—Spain, Italy, Chile, and Colombia—are responding to the growing burden of multimorbidity and frailty among older adults, integrating quantitative demographic indicators, policy document review, and expert interviews within a convergent mixed-methods design. The central finding is a consistent misalignment between the epidemiological burden of multimorbidity and frailty, the stated ambition of national policy frameworks, and the degree of operational implementation across all four countries.

The quantitative results confirm a marked demographic contrast between the two country groups. Chile and Colombia recorded CAGR of their aging indices exceeding 4%, with estimated doubling times of approximately 17 years, compared with 1.96% and 1.39% for Italy and Spain, respectively (doubling times of 36–50 years). These figures are consistent with projections from the United Nations Population Division (2022) and with regional analyses documenting Latin America as one of the fastest-aging regions globally (Gutiérrez Robledo et al., 2022). Importantly, accelerating trends were observed in three of the four countries during 2020–2024, suggesting that demographic pressure on health systems will intensify in the near term. The combination of rapid demographic transition and persistently high avoidable mortality in Latin American settings (247 per 100,000 in Chile and 328 in Colombia, compared with 146 in Italy and 163 in Spain) highlights the urgency of strengthening system preparedness before the burden of multimorbidity and frailty reaches levels already observed in Southern Europe.

Despite these differences in demographic velocity, the policy document review revealed that all four countries have established formal frameworks addressing population aging, multimorbidity, or frailty. However, the translation of these frameworks into operational practice was uneven across and within countries, a finding that emerged consistently from both documentary analysis and expert interviews. Spain and Italy have developed more explicit policy recognition of frailty as a clinical and care construct, supported by screening protocols and integration mechanisms, yet continue to experience fragmented implementation and territorial heterogeneity. Chile and Colombia have articulated broad strategic responses centered on functional capacity and social determinants, but these remain largely pilot-based, with limited standardization and scale. This observation is consistent with evidence from the 2025 Lancet Commission on Frailty, which concluded that frailty remains poorly understood among policy-makers globally and that, without adequate causal frameworks, preventive strategies cannot be effectively designed (Dent, Clegg, et al., 2025). Similarly, a recent New England Journal of Medicine review emphasized the need for pragmatic, system-level frailty assessment to inform clinical management, particularly in primary care settings where the benefit of routine screening remains to be established (Kim & Rockwood, 2024).

The qualitative findings further illuminate the nature of the implementation gap. Expert informants across countries described persistent barriers including the continued dominance of disease-oriented workflows, limited community-based care capacity, workforce constraints, and financing arrangements that prioritize institutional over community care. These barriers are consistent with those documented in recent reviews of integrated care for older adults (Briggs et al., 2018; Dlima et al., 2024) and with evidence from the WHO’s ICOPE framework, which identifies the integration of health and social services in primary care as a prerequisite for effective frailty management (WHO, 2019). Conversely, enabling factors reported by participants—trained multidisciplinary teams, shared assessment criteria, information systems supporting continuity of care, and strong ministerial stewardship—align with conditions identified in European and international studies as critical for scaling integrated care beyond isolated initiatives (Hendry et al., 2025; Wachholz et al., 2024).

A notable finding was the heterogeneity in how frailty is conceptualized and assessed across the four countries. Spain has adopted the Fried phenotype in primary care settings linked to hospital utilization patterns; Italy has embedded frailty within multidimensional biopsychosocial assessment frameworks connected to integrated care plans; Chile uses functional assessment as a proxy for vulnerability without formal operationalization of frailty; and Colombia employs the FRAIL scale in pilot settings while centering clinical decisions on functional capacity. This diversity reflects both context-specific adaptation and the absence of international consensus on a single frailty assessment standard (Hoogendijk et al., 2019; Dent, Martin, et al., 2019). It also highlights a shared challenge: connecting frailty identification to actionable care pathways. Recent evidence from primary care settings in Italy has demonstrated the feasibility of two-step frailty screening approaches that link identification to integrated care activation, offering a potentially transferable model for other contexts (Milani et al., 2025).

From a comparative perspective, the findings suggest that Chile and Colombia face a narrow window of opportunity to adapt their health systems before the burden of multimorbidity and frailty reaches the levels already documented in Southern Europe. Recent evidence indicates that health systems which delay investment in primary care, community services, and long-term care integration tend to replicate the same implementation gaps observed in more aged settings, including excessive hospital use and fragmented care trajectories (Beard et al., 2016; Lloyd-Sherlock et al., 2016). At the same time, the European experience demonstrates that policy maturity alone does not guarantee operational success: Spain and Italy, despite decades of policy development, continue to confront territorial heterogeneity, workforce shortages, and insufficient community infrastructure. This suggests that governance capacity, financing alignment, and workforce development are at least as important as formal policy recognition in determining system preparedness (Dlima et al., 2024; Gutiérrez Robledo et al., 2022). Recent analyses of primary care policy investments in Latin America further underscore the role of sustained financing and institutional governance in enabling effective responses to chronic care needs (Massuda et al., 2025).

A cross-cutting theme that emerged from the analysis is the importance of the social dimension of aging for health system responses. Spain and Italy have more explicitly incorporated health–social care integration into policy frameworks, recognizing that frailty risk emerges from the interaction between functional decline, multimorbidity, and social vulnerability. In Chile and Colombia, although the importance of social determinants is acknowledged in policy documents, social care remains weakly integrated into routine health service delivery. This observation aligns with evidence from Nature Medicine documenting that social disparities, cardiometabolic disease, and mental health are the principal predictors of healthy aging in Latin American populations (Santamaria-Garcia et al., 2023), and supports the argument that effective multimorbidity and frailty management requires attention to social as well as clinical determinants.

This study has several strengths. The convergent mixed-methods design enabled triangulation across quantitative indicators, policy documents, and qualitative expert evidence, providing a more comprehensive understanding of policy-implementation gaps than any single method could achieve. The inclusion of countries at different stages of demographic aging and system maturity enhances the analytical value of the comparison and supports the identification of transferable lessons. The use of national health systems as units of analysis is methodologically appropriate for cross-country policy comparison (Creswell & Plano Clark, 2018).

Several limitations should be acknowledged. First, the qualitative component was based on six expert informants, which may not fully capture within-country heterogeneity, particularly in decentralized health systems such as Spain and Italy. However, the sample size is consistent with the study’s design rationale, which treated each national health system as the unit of analysis and prioritized focused comparative insight over broad stakeholder representation. Purposive sampling of senior informants, combined with triangulation across data sources, enhances the analytic rigor of the qualitative component (Malterud et al., 2016; Palinkas et al., 2015). Second, cross-country comparisons were constrained by differences in data availability, definitions of multimorbidity and frailty, and the use of administrative versus population-based sources, which limit direct comparability of prevalence estimates. The absence of standardized long-term care data for Chile and Colombia reflects genuine data gaps in these settings, not analytical oversight. Third, the study focuses on documented strategies and reported experiences rather than measured outcomes, limiting causal inference regarding the effectiveness of specific interventions or policies. Fourth, the targeted literature review, while appropriate for the study’s comparative and policy-oriented objectives, did not follow a systematic review protocol, and relevant sources may have been missed. Despite these limitations, the integrated mixed-methods design and structured comparative framework provide a robust and policy-relevant assessment of current responses to multimorbidity and frailty in aging societies.

## Conclusions

This comparative mixed-methods analysis demonstrates that multimorbidity and frailty represent central and growing challenges for aging societies across diverse health system contexts, irrespective of their current stage in the demographic transition. The findings reveal a consistent pattern in which epidemiological pressure and policy ambition are not yet matched by operational implementation, particularly in relation to integrated, function-oriented care.

Spain and Italy have more explicitly incorporated the social dimension of aging into health and social care policies, recognizing that frailty risk emerges from the interaction between functional decline, multimorbidity, and social vulnerability. In these settings, the integration of health and social services is acknowledged as a core component of frailty prevention and care, although implementation remains uneven across territories. In Chile and Colombia, comprehensive aging policies are in place and the importance of social determinants is recognized, yet frailty is less clearly operationalized as a clinical and care construct: approaches remain largely implicit, centered on functional status or chronic disease management, with limited use of standardized assessment and weak integration of social care into routine health service delivery.

The main challenge lies not in the absence of policy frameworks, but in their effective implementation. Across all four countries, qualitative evidence underscores the importance of care models centered on functional capacity, multidisciplinary teams, coordination between health and social services, and information systems that support continuity of care. Countries facing similar demographic and system pressures may benefit from prioritizing early investment in primary and community care, explicit operational guidance for frailty and multimorbidity management, and governance and financing mechanisms aligned with integrated, person-centered models.

From a practice perspective, strengthening responses to multimorbidity and frailty requires embedding existing policy commitments into routine care processes. Practical priorities include incorporating systematic, function-oriented frailty assessment into primary care workflows; establishing clear referral and coordination pathways between health and social services; and supporting multidisciplinary teams with defined roles and protected time for care planning and follow-up. Scaling effective pilot initiatives will depend on simplifying operational guidance, aligning information systems to support patient identification and continuity of care, and ensuring that financing mechanisms favor community- and home-based services over episodic, institution- centered care.

Future research should examine the long-term effectiveness of integrated care models for older adults with multimorbidity and frailty in Latin American settings, where accelerated aging dynamics and system fragmentation create particular challenges. Longitudinal studies tracking both health outcomes and implementation processes across different levels of governance would help identify the conditions under which policy frameworks translate into sustained improvements in care. Comparative work incorporating a broader range of countries and health system typologies could further strengthen the evidence base for context-adapted responses to population aging.

## Data Availability

The data underlying the findings of this study consist of publicly available documents and qualitative interview data. The public documents analyzed are cited within the manuscript and are available from their original sources. The qualitative interview data are not publicly available because they contain information that could compromise participant confidentiality and because participants did not provide consent for public sharing of the interview transcripts. De-identified excerpts supporting the findings are included in the manuscript. Additional de-identified information may be made available by the corresponding author upon reasonable request, subject to ethical considerations and applicable institutional requirements.

## Abbreviations

ADL: Activities of Daily Living
CAGR: Compound Annual Growth Rate
COPD: Chronic Obstructive Pulmonary Disease
ECICEP: Estrategia de Cuidado Integral Centrado en las Personas
ENS: National Health Survey (Chile)
ENSE: National Health Survey of Spain
FIASO: Federazione Italiana Aziende Sanitarie e Ospedaliere
IADL: Instrumental Activities of Daily Living
INE: Instituto Nacional de Estadística (Spain)
ISTAT: Istituto Nazionale di Statistica
LTC: Long-Term Care
MACEP: Multimorbidity Patient-Centered Care Model
MINSAL: Ministry of Health of Chile
OECD: Organization for Economic Co-operation and Development
PAHO: Pan American Health Organization
PaRIS: Patient-Reported Indicator Surveys
WHO: World Health Organization.

## Declarations

### Ethical Approval

This study was conducted in accordance with the principles of the Declaration of Helsinki and with international ethical standards for qualitative health policy research. Ethical approval was granted by the Research Ethics Committee of the Fundación Santa Fe de Bogotá (approval communication: CCEI-16596-2024, issued 27 June 2024).

## Acknowledgments

The authors wish to acknowledge the institutions and experts from Spain, Italy, Chile, and Colombia who contributed to this study through their participation in interviews, field visits, and provision of technical and contextual information. Their insights and experience were essential for understanding national approaches to multimorbidity and frailty and for interpreting policy frameworks, implementation processes, and service delivery practices across diverse health system contexts. We are particularly grateful to the professionals working in health and social care services, public institutions, and academic settings who generously shared their time and expertise, thereby strengthening the depth and relevance of this comparative analysis.

## Consent to Participate

All expert interview participants provided verbal informed consent prior to participation. Interviews were conducted virtually, and all participants agreed to participate and to be audio-recorded.

## Consent for Publication

Not applicable. The manuscript contains no individual personal data, images, or identifiable participant information. All qualitative data are presented in aggregate and fully de-identified form.

## Availability of Data and Materials

The quantitative data analyzed in this study are derived from publicly available international databases and national statistical repositories, which are cited in the article. The qualitative data (expert interview transcripts) generated during the study are not publicly available due to confidentiality restrictions. De-identified excerpts are available from the corresponding author upon reasonable request.

## Authors’ Contributions

OV contributed to conceptualization, methodology, formal analysis, investigation, data curation, writing – original draft, writing – review and editing, visualization, and project administration.

OB, CD, MM, PZ, NP, LFGF, and GL contributed to conceptualization and review and editing. SLG contributed to formal analysis and writing – review and editing. All authors reviewed and approved the final version of the manuscript.

